# Continuous pain report demonstrates time delay of pain ratings in Fibromyalgia

**DOI:** 10.1101/2020.12.28.20248780

**Authors:** Anna Y Kharko, Stephen D Hall, Paul L Furlong, Matthew E Roser

## Abstract

**Background:** Enhanced temporal summation (TS), measured through self-reported pain ratings, has been interpreted as indicative of central sensitisation in fibromyalgia. Greater TS in patients, however, has not been universally observed. It is also unclear whether increased pain report maintains beyond the TS period.

**Methods:** In this study, we measured TS through continuously reported pain ratings. Fibromyalgia-diagnosed patients (*n* = 17) and matched pain-free controls (*n* = 13) rated painful transcutaneous electrical stimulation of various intensity levels in 18 one-minute-long blocks. Pain was rated on a 101-point visual analogue scale. The resulting continuous response was divided into TS (< 15s) and adaptation (15 – 60s) periods. Average pain values were extracted for each period alongside the timing of key events such as maximal pain ratings. The difference in temporal summation and adaptation measures between fibromyalgia and control participants was analysed using mixed-effects modelling.

**Results:** The average pain ratings for TS and adaptation periods were not significantly associated with fibromyalgia diagnosis but were with stimulation intensity. The same was true for the magnitude of the maximal rating during TS and the slope leading to that peak rating. The presence of fibromyalgia, however, did predict the time of the maximal TS rating, as well as the value and the time of the maximal adaptation rating.

**Conclusions:** Our study did not find homogeneously increased TS pain ratings. Instead, by utilising continuous pain data we demonstrate for the first time that the time of TS peak rating, as well as the magnitude and time of adaptation peak rating are linked to fibromyalgia diagnosis.

## INTRODUCTION

Fibromyalgia (FM) is a chronic widespread pain condition of unknown aetiology, associated with disrupted sleep, fatigue and mild cognitive disturbances; resulting in high emotional burden (Clauw, 2014). The high prevalence, 2% to 5% of the population (Queiroz, 2013), and limited long-term success of symptom management (Häuser et al., 2014), has put pressure on understanding the mechanisms that underlie pain processing in FM. Allodynia and hyperalgesia, the key markers of FM pain (Sluka & Clauw, 2016), have been attributed to deficient pain inhibition in the central nervous system (CNS) (Meeus & Nijs, 2007), a phenomenon known as central sensitisation. In psychophysical research, it has been studied through temporal summation (TS).

TS is the increase in reported pain in response to unremitting stimulation at a fixed frequency (Arendt-Nielsen et al., 2000). It is considered the behavioural counterpart of wind-up (WU), a central spinal mechanism where an increased excitation of dorsal horn neurons is observed following repeated engagement of C-fibres (Gebhart & Schmidt, 2013). While TS and WU are part of healthy nociception, both have been reported to be augmented in chronic pain conditions, such as FM (Staud, 2012).

The majority of observations on TS in FM come from a series of studies began by Staud and Price (Staud et al., 2001). They have reported that FM patients require lesser stimulation intensity than pain-free controls to produce similar TS pain ratings (Staud et al., 2003). Further, maintenance of TS required less stimulation in the FM cohort than control (Staud et al., 2004). Importantly, this pattern of increased TS in FM has been demonstrated across several stimulation modalities (Graven-Nielsen et al., 2000; Staud et al., 2003, 2014).

Greater TS in FM, however, is not a universally-observed phenomenon. Several studies report mixed results, in which increased TS in patients was contingent on stimulation location or modality (for review see O’Brien (2018). One study failed to find any deviancy in the clinical cohort (Lim et al., 2016). Part of this literature heterogeneity may stem from varying methods of TS elicitation and measurement. The conventional approach dictates deriving a measure based on comparison between a single pain rating and a later rating following repeated stimulation at a frequency that evokes TS (Lim et al., 2016; Staud et al., 2001). Such a low rate of pain sampling provides a coarse measure of the *temporal* aspect of TS and does not show whether the enhanced pain perception maintains beyond the period of TS. The following period, here termed adaptation, is also a marker of WU and can be used to characterise central sensitisation (Graven-Nielsen et al., 2000) but has not yet been adopted in FM.

The conventional approach has limited utility. Firstly, the use of singular pain ratings does not capture the possible development of pain perception across a fixed interval. Secondly, the reporting of a single value from a point in time is likely subject to high measurement error and individual variation. Thirdly, the serial processes of summation and adaptation are likely to be conflated by use of a fixed measurement point and the temporal envelope of these processes overlooked.

Here, we describe the collection of continuous pain ratings, to address the limitations of single fixed pain reports. Continuous pain ratings, concurrent to stimulation, have been found to reliably reflect acute pain perception (Boormans et al., 2009; Wijk et al., 2013). In FM research, one study used continuous pain report but did not analyse the time property of the gathered pain data (Potvin et al., 2012). The extraction of key timepoints from TS and adaptation for comprehensive analysis of acute pain processing in the presence of FM is yet to be done.

To address this, we carried out a study with FM patients and pain-free controls using transcutaneous electrical stimulation (TES), a well-established method for eliciting TS (Arendt-Nielsen et al., 2000). We applied tonic single-pulse TES at individual-derived intensity levels and asked participants to rate their pain continuously on an automated visual analogue scale (VAS). We anticipated that time, a new property of the extracted data, will be the key measure to characterise TS in FM.

## METHODS

### 2.1. Participants

Participants were recruited on the basis of several strict eligibility criteria (see Table 1). Ethical permission was granted by the School of Psychology, University of Plymouth (17/18-890).

**Table 1.**
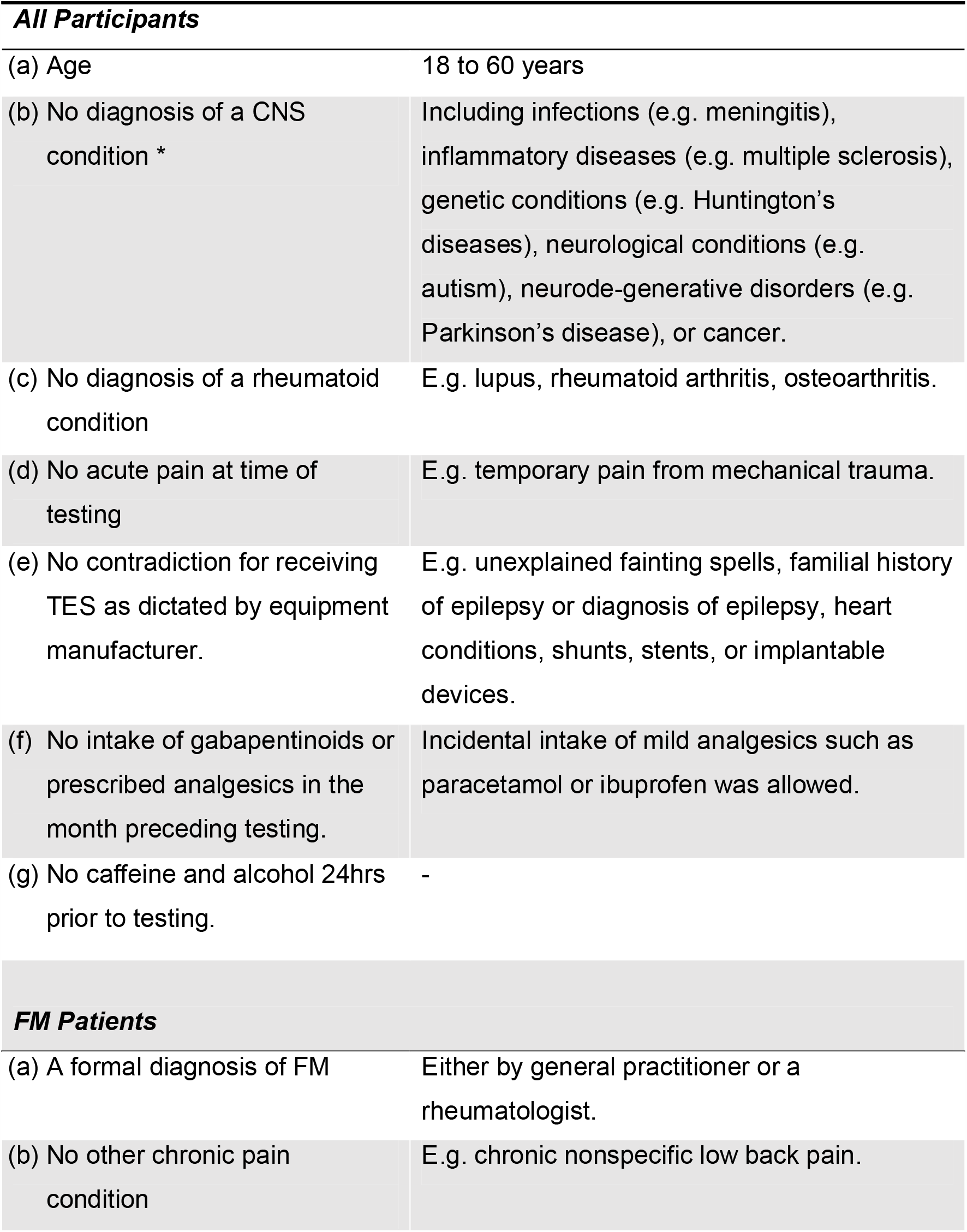
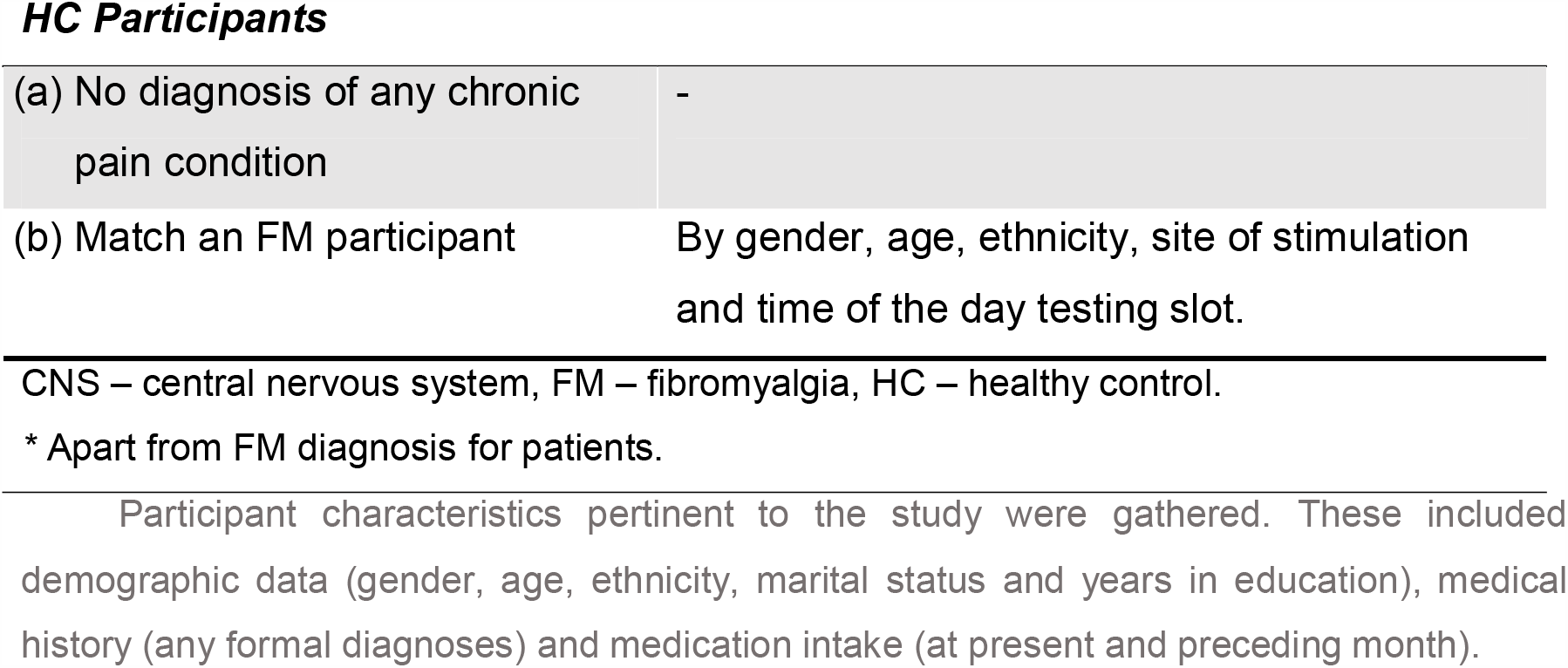
Eligibility Requirements for Participation

| **All Participants** |  |
| --- | --- |
| (a) Age | 18 to 60 years |
| (b) No diagnosis of a CNS condition * | Including infections (e.g. meningitis), inflammatory diseases (e.g. multiple sclerosis), genetic conditions (e.g. Huntington's diseases), neurological conditions (e.g. autism), neurodegenerative disorders (e.g. Parkinson's disease), or cancer. |
| (c) No diagnosis of a rheumatoid condition | E.g. lupus, rheumatoid arthritis, osteoarthritis. |
| (d) No acute pain at time of testing | E.g. temporary pain from mechanical trauma. |
| (e) No contradiction for receiving TES as dictated by equipment manufacturer. | E.g. unexplained fainting spells, familial history of epilepsy or diagnosis of epilepsy, heart conditions, shunts, stents, or implantable devices. |
| (f) No intake of gabapentinoids or prescribed analgesics in the month preceding testing. | Incidental intake of mild analgesics such as paracetamol or ibuprofen was allowed. |
| (g) No caffeine and alcohol 24hrs prior to testing. | - |
| **FM Patients** |  |
| (a) A formal diagnosis of FM | Either by general practitioner or a rheumatologist. |
| (b) No other chronic pain condition | E.g. chronic nonspecific low back pain. |
| **HC Participants** |  |
| (a) No diagnosis of any chronic pain condition | - |
| (b) Match an FM participant | By gender, age, ethnicity, site of stimulation and time of the day testing slot. |

CNS – central nervous system, FM – fibromyalgia, HC – healthy control.
\* Apart from FM diagnosis for patients.

### 2.2. Psychological Testing

Presence or absence of FM was confirmed on the day of testing through the American College of Rheumatology Criteria from 1990, ACR ‘90, and 2010, ACR ‘10 (Wolfe et al., 1990, 2010). The latter were scored considering the redactions from 2016 (Wolfe et al., 2016). Recent pain history was assessed through the Short-Form McGill Pain Questionnaire, SF-MPQ (Melzack, 1987).

### 2.3. Psychophysical Testing

All psychophysical testing was performed on the skin over the sural nerve at lateral border of tendo-Achilles. The testing setup can be seen in Figure 1. Body side was counterbalanced between participants. The area was to be free of skin damage, tattoos, and hair. Prior to placing electrodes, the stimulation site was cleaned with an abrasive gel followed by 70% isopropyl alcohol. Two disposable 2 cm self-adhesive disk electrodes were then positioned with inter-electrode distance of 3 cm. Impedance was checked by D175 impedance meter (Digimeter Ltd., United Kingdom) and was maintained below 50 kohm.

**Figure 1.**
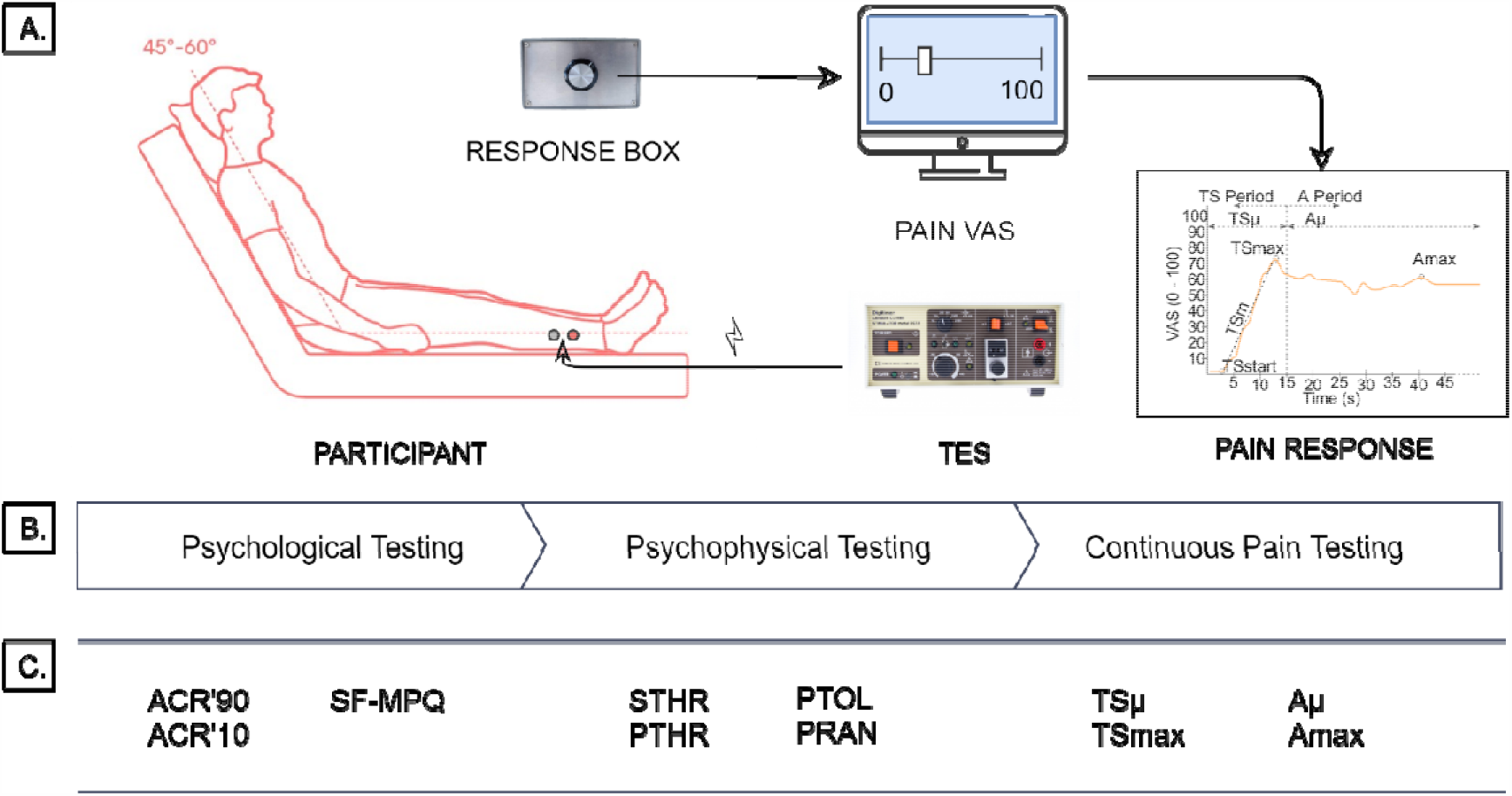
Experimental Setup. A. Equipment setup; B. Study stages; C. Acquired measures at each study stage. VAS – visual analogue scale; TES – transcutaneous electrical stimulator; ACR’90/’10 – American College of Rheumatology Criteria for fibromyalgia diagnosis from 1990 and 2010; SF-MPQ – Short-Form McGill Pain Questionnaire; STHR – sensory threshold; PTHR – pain threshold; PTOL – pain tolerance; PRAN – pain range; TS/Aμ – average pain rating during temporal summation/adaptation; TS/Amax – maximal pain rating during TS/A; Amin – minimal pain rating during A. Stimulator image is supplied by manufacturer (Digitimer Ltd., UK) and sitting position image from Dimensions.Guide. Reproduced from respective sources with permission.

#### Stimulation Parameters

Stimulation was delivered by a constant current stimulator DS7AH Digitimer (Digitimer Ltd., UK). The equipment was operated semi-automatically by the same experimenter: a computer maintained the frequency while the experimenter adjusted the intensity. Stimulation was a single-square wave pulse of 500 µs duration, with maximum of 400V for the output.

#### Psychophysical Measures

To derive participant-specific stimulation levels, static quantitative sensory testing (QST) was first performed. Three psychophysical measures were collected: innocuous stimulus detection (ISD), pain threshold (PTHR), and pain tolerance (PTOL). ISD was the first instance of any sensation at the site of stimulation. PTHR was the first instance of stimulation being perceived as painful. PTOL was the first instance of participant indicating that they no longer wish to experience the next stimulation level increase. At each point, participants described the sensation to ensure understanding of instructions. Static QST was performed three times and each measure was defined as the average from these.

At the start of stimulation, the current was set at 0 mA and then increases in 0.1 mA increments. Stimulation was delivered manually by experimenter at an approximate interval of 1 s to prevent adaptation. Maximal current intensity was predetermined to be 40 mA. If PTOL was not established prior to that, the stimulation was stopped. If more runs were left, the procedure was restarted. If during all runs the limit was reached, it was deemed that a participant’s pain tolerance was not established, and they were excluded from further participation.

At the end of the procedure a secondary psychophysical measure was derived: pain range (PRAN), which was determined as the difference between pain threshold and tolerance (PTOL – PTHR = PR).

### 2.4. Continuous Pain Testing

#### Stimulation Conditions

There were six stimulation levels 10%PRAN, 20%PRAN, 30%PRAN, 40%PRAN, 50%PRAN, and 60%PRAN, calculated as PTHR + (PRAN x %/100). Each stimulation intensity was delivered three times, resulting in 18 experimental blocks, presented in a computer-randomised order. Short breaks (no more than 1 min) were available between each block, with mandatory longer breaks after the 6^th^ and 12^th^ blocks (no more than 5 min).

#### Stimulation Parameters

During continuous pain testing, stimulation protocol was adjusted to elicit TS. The current remained as single-wave pulses with 500 µs duration and up to 50 V. Instead of manual stimuli delivery, a computer maintained a 2.5 Hz frequency. Current intensity was dictated by stimulation condition and was thus always within acceptable to the participant range. The experimenter was responsible for the manual adjustment of intensity and did so before each block, without the awareness of the participant.

#### Continuous Pain Rating

To continuously reassess pain, a pain visual analogue scale (VAS) was used. The scale ranged from ‘0’ to ‘100’, with major ticks at every tenth mark. On the scale ‘0’ was no pain, ‘1’ was minimal pain, and ‘100’ was the worst pain imaginable. Participants were instructed to constantly rate the experienced pain for the duration of the stimulation block. If other pain was present, which was the case for the clinical cohort, participants were to exclude it from the rating.

The rating was provided using a proprietary response device fitted with an Arduino Genuino (Arduino LLC) and a rotary button. By adjusting the rotation of the button, participants adjusted a sliding marker on the VAS. It is the position of that marker that was constantly resampled every 200 ms.

### 2.5. Analysis

Analysis was carried out in RStudio (v. 1.0.136).

Participant characteristics were summarised through average values and standard deviation (*SD*). Group differences were assessed through separate Student’s t-tests (two-tailed, with post-hoc Bonferroni correction) where appropriate.

#### Psychological Testing

Cumulative and subsection questionnaire scores were summarised as average values and SD. Student’s t-test (two-tailed) was used to compare cohorts on all forms.

#### Psychophysical Testing

STHR, PTHR and PTOL were calculated as the mean of each of the three recorded measurements. The average of each values was compared between groups through a Student’s t-test (two-tailed, with post-hoc Bonferroni correction). Group level mean, SD, and range were used to describe psychophysical values. The same procedure was repeated for stimulation conditions.

#### Continuous Pain Testing

To assess whether the presence of FM diagnosis was related to increased average and continuous pain ratings, key events and timepoints were extracted from the data for analysis. Figure 1, panel A) shows a prototypical pain response with measures of interest. First the response was broken into period of TS (0 to 15 s) and period of adaptation (15 s to 60 s). An average pain rating was calculated for each period (**TS**μ and **A**μ) to assess the overall pain rating for that period. Then maximal pain ratings were then extracted for each period (TSmax and Amax). Both value and time were recorded for each, resulting in the variables **TSmax-v, TSmax-t, Amax-v, Amax-t**. Lastly, for the TS period we calculated the slope from start of response to TSmax (**TSm**) to quantify the time needed to reach maximal rating during TS.

To analyse what factors predict the measures we used mixed-effect modelling. Separate models were created for each of the seven outcome variables. Predictors’ inclusion was predetermined based on relevancy to hypotheses. *Group* (FM vs HC) was entered as a fixed factor to determine whether the presence of a diagnosis is associated with an increase in pain measures. *Stimulation* (stimulation intensity in mA) was entered as a fixed factor to account for the mediating effects of stimulation intensity. And finally, *Participant* was entered as a random effect, to account for individual variance.

Model syntax was guided by protocol described elsewhere (Brysbaert & Stevens, 2018; Matuschek et al., 2017), suitable for minimising Type I error in small samples. Sample size was estimated through power simulations following an established method (Green & MacLeoud, 2016). A Kenward-Roger approximation was calculated for each fixed covariate, with the goal of observing ≥ 80% power for effect size .5.

## RESULTS

33 participants met all eligibility requirements. Three participants from the control cohort were unable to complete the experiment due to failure to establish PTOL, thus leaving a sample of 30 participants. 17 were FM participants and 13 were pain-free controls.

### 3.1. Participants & Psychological Testing

All participants were Caucasian females. No significant differences were found between samples on individual characteristics (see Table 2). Psychological testing did find that the FM sample experienced more pain due to their condition in the week preceding testing.

**Table 2.** Descriptive Statistics for Participants Samples & Psychological Testing

|  | *Mean (SD)* |  | *p-value* |
| --- | --- | --- | --- |
|  | *FM* | *HC* |  |
| Age (n years) | 35.35 (11.9) | 35.7 (12.84) | .941 |
| Education (n years) | 17.88 (2.23) | 18.46 (2.3) | .492 |
| Marital Status (n married) <sup>a</sup> | 9 (53%) | 4 (31%) | - |
| FM diagnosis duration (years) | 3.63 (3.13) | - | - |
| FM symptoms duration (years) | 10.04 (7.98) | - | - |
| Medication in the last 30 day <sup>*</sup> | 17 (100%) | 4 (31%) | - |
| Analgesics <sup>a b</sup> | 12 (71%) | 3 (18%) | - |
| Anticonvulsants <sup>a</sup> | 1 (6%) | - | - |
| Antidepressants <sup>a</sup> | 10 (59%) | 1 (8%) | - |
| ACR'90 |  |  |  |
| Widespread Pain | 7.76 (.75) | .15 (.38) | < .001 *** |
| Tender Points | 14.24 (2.49) | .31 (.63) | < .001 *** |
| ACR'10 |  |  |  |
| Widespread Pain Index | 13.88 (3.18) | 1.54 (1.44) | < .001 *** |
| Somatic Symptoms Severity | 7.65 (1.58) | 1.92 (2.22) | < .001 *** |
| Other Somatic Symptoms | 2.06 (.56) | 1.08 (.64) | < .001 *** |
| SF-MPQ |  |  |  |
| Pain Descriptors | 24.77 (7.4) | 2.85 (2.61) | < .001 *** |
| Visual Analog Scale | 6.65 (1.37) | 1.15 (.8) | < .001 *** |
| Present Pain Intensity | 3 (1.23) | - | - |

SD – standard deviation, FM – fibromyalgia group, HC – healthy control group. P-values were calculated using Student's t-test. Post-hoc Bonferroni correction indicated corrected p-value of < .006.
<sup>a</sup> Number and percentage calculated.
<sup>b</sup> Analgesics included mild over the counter medication.
\*\*\* Statistically significant.

### 3.2. Psychophysical Testing

No significant group differences were observed for STHR and PTHR (see Table 3). Both PTOL and PRAN were significantly higher in the clinical sample, just as all stimulation levels.

**Table 3.** Average Values (mA) for Psychophysical Measures & Stimulation Levels

|  | *FM* |  | *HC* |  | *p-value* |
| --- | --- | --- | --- | --- | --- |
|  | *Mean (SD)* | *Range* | *Mean (SD)* | *Range* |  |
| STHR | 1.38 (0.49) | .73 – 2.3 | 1.5 (0.33) | 1 – 1.97 | .427 |
| PTHR | 4.71 (2.83) | 1.97 – 13.17 | 6.57 (3.62) | 2.43 – 13.9 | .124 |
| PTOL | 11.82 (7.85) | 5.3 – 34.33 | 22.14 (9.75) | 9.33 – 38.9 | .003 *** |
| PRAN | 7.11 (5.7) | 2.53 – 24.5 | 15.57 (8.22) | 5.6 – 27.5 | .002 *** |
| 10% PRAN | 5.42 (3.23) | 2.3 – 14.69 | 8.13 (3.89) | 3.12 – 16.37 | .047 |
| 20% PRAN | 6.13 (3.67) | 2.7 – 16.22 | 9.68 (4.32) | 3.81 – 18.87 | .022 |
| 30% PRAN | 6.84 (4.15) | 3 – 17.75 | 11.24 (4.84) | 4.5 – 21.37 | .012 |
| 40% PRAN | 7.55 (4.65) | 3.3 – 19.63 | 12.8 (5.44) | 5.19 – 23.87 | .008 |
| 50% PRAN | 8.27 (5.16) | 3.7 – 22.08 | 14.35 (6.09) | 5.88 – 26.37 | .006 |
| 60%PRAN | 8.98 (5.69) | 4 – 24.53 | 15.91 (6.78) | 6.57 – 28.87 | .005 *** |

SD – standard deviation, FM – fibromyalgia group, HC – healthy controls group, STHR – sensory threshold, PTHR – pain threshold, PTOL – pain tolerance, PRAN – pain range. P-values were calculated using Student's t-test. Post-hoc Bonferroni correction indicated corrected p-value of < .005.
\*\*\* Statistically significant.

### 3.3. Continuous Pain Testing

The best mixed-fixed effects model for all measures had the following formula:

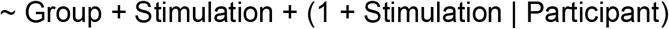

A full model summary can be found in Supplementary Material 1. Analysis of **TS**μ showed that it is significantly associated with Stimulation (*Coeff*. = 5.56, *SE* = 6.52, *t* = 7.14, *p* < .001) but not Group (*Coeff*. = 10.64, *SE* = 6.52, *t* = 1.63, *p* = .110), indicating that an increase in stimulus intensity was reflected in the average rating for the period, unlike the presence of diagnosis. Marginal means estimated based on that model show a trend for increased pain rating in FM group (see Table 4). This trend, however, was not statistically significant, a pattern that was repeated in the subsequent analyses of TS pain measures.

**Table 4.** Estimated Marginal Means.

| Group | EMM | SE | 95% CI |  |
| --- | --- | --- | --- | --- |
|  |  |  | Lower | Upper |
| TS $\mu$ | | | | |
| HC | 24.12 | (6.49) | 11.03 | 37.22 |
| FM | 34.76 | (5.86) | 22.94 | 46.59 |
| TSmax.v |  |  |  |  |
| HC | 36.47 | (9.61) | 17.09 | 55.86 |
| FM | 50.51 | (8.69) | 32.98 | 68.03 |
| TSmax.t * |  |  |  |  |
| HC | 7.38 | (.98) | 5.39 | 9.37 |
| FM | 10.56 | (.91) | 8.72 | 12.39 |
| TSm |  |  |  |  |
| HC | 6.36 | (.79) | 4.74 | 7.97 |
| FM | 6.76 | (.71) | 5.32 | 8.2 |
| Aμ | | | | |
| HC | 30.48 | (9.61) | 11.05 | 49.91 |
| FM | 50.52 | (8.6) | 33.15 | 67.88 |
| Amax.v * |  |  |  |  |
| HC | 29.25 | (9.3) | 10.39 | 48.12 |
| FM | 59.05 | (8.32) | 42.22 | 75.88 |
| Amax.t * |  |  |  |  |
| HC | 24.33 | (3.31) | 17.58 | 31.07 |
| FM | 36.81 | (3.05) | 30.66 | 42.97 |

EMM – estimated marginal means, SE – standard error, CI – confidence interval.
\* Significant ( $p < .05$ ) group difference established in respective linear mixed-effects models.

The magnitude of the maximal TS pain rating (**TSmax**.**v**) was similarly predicted by Stimulation (*Coeff*. = 7.81, *SE* = 1.08, *t* = 7.22, *p* < .001) but not the Group (*Coeff*. = 14.03, *SE* = 9.53, *t* = 1.47, *p* = .152). The time when the maximal rating was made (**TSmax**.**t**), however, was significantly associated both with Stimulation (*Coeff*. = .51, *SE* = .09, *t* = 5.46, *p* < .001) and Group (*Coeff*. = 3.17, *SE* = 1.23, *t* = 2.58, *p* = .016). Increase in stimulation slightly increased the time to the peak rating. Separately, belonging to the FM cohort was associated with an additional almost 3 s delay (see Table 4).

The slope from the start of the response to the maximal pain rating (**TSm**) was not related to Stimulation (*Coeff*. = .14, *SE* = .08, *t* = 1.8, *p* = .089) or Group (*Coeff*. = .41, *SE* = .98, *t* = .42, *p* = .681), suggesting that the slope did not differ on basis of stimulus intensity nor the presence of FM diagnosis.

The average pain rating during the period of adaptation (**A**μ) significantly increased with Stimulation (*Coeff*. =8.15, *SE* = .98, *t* = 8.29, *p* < .001) but not Group (*Coeff*. = 20.04, *SE* = 10.6, *t* = 1.89, *p* = .069). As with TSμ, the trend exhibited by FM patients for higher average pain rating for the period did not reach statistical significance.

In contrast, the maximal rating made during adaptation (**Amax**.**v**) was sensitive to both stimulation and presence of diagnosis. An increase of Stimulation intensity was associated with an increase of reported pain (*Coeff*. = 7.96, *SE* = .97, *t* = 8.23, *p* < .001). The FM Group additionally produced significantly higher maximal pain ratings during adaptation (*Coeff*. = 29.8, *SE* = 10.87, *t* = 2.74, *p* = .011). The same was true for the time the peak rating was made (**Amax**.**t**). Increased Stimulation (*Coeff*. = 1.26, *SE* = .37, *t* = 3.41, *p* = .006) and belonging to the FM group (*Coeff*. = 12.49, *SE* = 4.1, *t* = 3.04, *p* = .005) predicted a delay in reaching that maximal rating.

## DISCUSSION

Despite a large body of research reporting augmented TS in FM (O’Brien et al., 2018), several studies have failed to consistently achieve the same results (Lim et al., 2016; Potvin et al., 2012; Staud et al., 2008). To address this discrepancy, we adopted a different pain measurement approach, in which pain perception was assessed through continuously gathered pain ratings. Using it, we were able to analyse not only the value of a given pain rating but also the time it was made.

We found that FM was significantly associated with delays in reaching peak pain ratings during the periods of TS and adaptation. In contrast, only the magnitude of the maximal peak rating during adaptation differed significantly between cohorts.

The value of the peak TS rating was not significantly associated with diagnosis, and neither was the slope to that peak. Average pain rating during TS and adaptation were also not found to be significantly different between participant groups. The best mixed-effects model for each measure included stimulation intensity as a factor. Apart from the TS slope, all measures were found to be predicted by it. An increase in stimulus intensity was mirrored by an increase in average pain ratings during TS and adaptation, as well as magnitude and time of maximal ratings. Together, the findings show that continuous pain report not only enabled the extraction of a new variable property but that it was this temporal property that was consistently associated with the presence of FM diagnosis.

Although using the maximal pain rating in response to TS eliciting stimulation has been the conventional approach to characterising central sensitisation in FM (Staud et al., 2001), our study suggests that this may be only one of several markers of deficient pain modulation. In previous studies, pain ratings were collected at predetermined timepoints following a prolonged stimulation needed to elicit TS (Staud et al., 2001). This both limited the measurement window and the number of observations. In our study, we purposefully extended the pain rating collection time. This gave us sufficient time to measure the TS part of the response, as well as to observe how participants adapt to pain post-TS. Let us consider both in succession. First, the extended record enabled the flexible extraction of maximal TS rating. We propose that it was this key change that allowed us to observe significant group differences in the TS period. It may be that previous research, which was not successful in finding augmented TS, measured peak TS rating too early, as our results indicate that FM patients were slow in reaching their peak rating. That delay is of particular importance when considered together with analysis of the later adaptation period.

Extending data collection past the TS period was the second major advancement of our study. The conventional focus on TS as a method of quantifying centrally dysregulated pain modulation in FM had demotivated further inspection into *how* participants adapt to prolonged pain. This is different to studies where aftersensations to TS-inducing pain stimulation were studied (Banic et al., 2004). Here, we were interested to observe whether FM participants continue to rate their pain increasingly high, thus indicating sensitisation, or whether they would slowly begin to habituate to it, evident in reduced pain ratings. The finding that maximal adaptation rating was not only higher but reached later by the FM group suggests that these participants continued to sensitise to the stimulation. The yet again delayed peak further supports the idea of disrupted pain inhibition under FM (Sluka & Clauw, 2016).

Our study also agreed with previous research that hyperalgesia is integral to the FM pain profile (Nielsen & Henriksson, 2007). We chose to apply individually-derived stimulation conditions in order to demonstrate the differences in sensitivity between the participant cohorts. Despite the stimulation levels being calculated so that comparable pain ratings are be observed between participants, this was not found. For example, the 10% PRAN condition should have elicited pain ratings around the 10th mark on the VAS, regardless of which group the participant belonged to. As could be seen in Figure 3, however, patients consistently provided ratings higher than their pain-free counterparts. Further, the average stimulation values were lower in the FM group, yet they still rated the evoked pain as higher than the control. Together, this pattern of results suggests that hyperalgesia, a key marker of central sensitisation, is present in FM.

**Figure 3.**
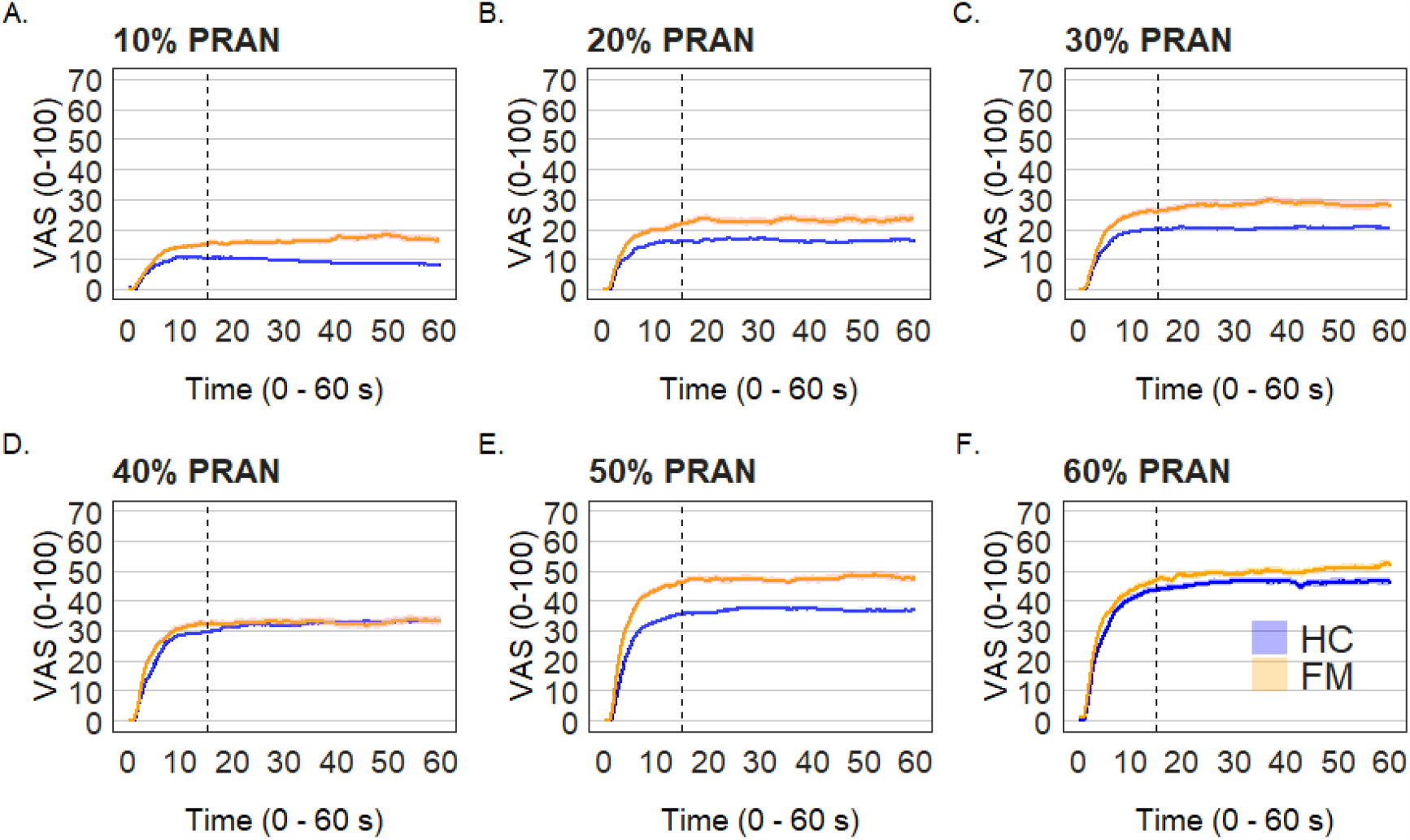
Average Trajectory of Continuous Pain Ratings per Stimulation Condition. VAS – visual analogue scale, PRAN – pain range; HC – healthy control; FM – fibromyalgia.

It is interesting that HC participants produced consistently lower ratings than those expected for the respective stimulation condition. Perhaps the trend is partially attributable to habituation. The ability to adjust to continuous mild nociceptive input is part of the CNS regulatory mechanisms. The failure to see a similar effect in FM participants further supports the theory of the CNS origin of the syndrome (Nielsen & Henriksson, 2007).

### Limitations & Future Directions

Unlike other studies (Staud et al., 2005), we only examined continuous pain report in response to TES. Research has shown that a comprehensive pain profile can only be achieved when testing pain perception through multiple stimulus modalities (Hastie et al., 2005). In FM, employing a multidimensional pain testing protocol with continuous pain ratings would further clarify the role of time in the augmented development of TS and adaptation. Further, the focus of the current study were only behavioural markers of central sensitisation. It remains unclear how does the subjective pain report connect to the physiological response. Although, TS and adaptation are assumed to reflect underlying WU (Graven-Nielsen et al., 2000), psychological modulators such as stress have been theorised to mediate self-reported pain at supraspinal level (Crettaz et al., 2013). A concurrent measurement of both TS and WU is not only plausible due the common stimulation protocol but would also be useful in the clarification of their relationship and allow for further investigation of psychological mediating factors. Lastly, while the benefits of individually derived stimulation levels were evident, they do complicate data interpretation. Here we calculated stimulation conditions using an individual’s pain range, which led us to deliver vastly different stimulation between cohorts. The alternative approach where predetermined values are used may be adopted instead.

## Conclusion

Continuous pain ratings of TES were simple to implement while rich in produced data. The newly extracted time property of the maximal pain ratings made during TS and adaptation were found to be reliable measures of differentiation between FM-diagnosed and pain-free cohorts. Analysis of the later pain response period, adaptation, was also beneficial for the characterisation of central sensitisation in FM, and should be analysed alongside of TS in future investigations.

## Data Availability

Data is available upon request from the corresponding author.

## Acknowledgments

The authors thank Carol Hambly, the organiser of a local Fibromyalgia Support Group for her support in participant recruitment.

