## Supplementary Material 1 for "Continuous pain report demonstrates time delay of pain ratings in Fibromyalgia"

Supplementary Material 1. Model Summaries

Table 1. Model Summary for **TSavg**

|  | *Coeff.* | *(SE)* | *t-value* | *p-value* |
| --- | --- | --- | --- | --- |
| *Fixed effects* | | | |  |
| Intercept | 27.47 | (6.13) | 4.48 | < .001 |
| FM Group | 10.64 | (6.52) | 1.63 | .110 |
| Stimulation | 5.56 | (.78) | 7.14 | < .001 |
|  | *σ^2^* | *(SD)* |  |  |
| *Random effects* | | | |  |
| Participant | 561.7 | (23.7) | - | - |
| Participant x Stimulation | 14.9 | (3.86) | - | - |
| Linear mixed-effects model was fit by maximum likelihood, t-tests were calculated automatically using Satterthwaite approximations to degrees of freedom.  Coeff./σ^2^ – estimated coefficient/variance, SE/SD – estimated standard error/deviation. | | | | |

Table 2. Model Summary for **TSmax.v**

|  | *Coeff.* | *(SE)* | *t-value* | *p-value* |
| --- | --- | --- | --- | --- |
| *Fixed effects* | | | |  |
| Intercept | 36.08 | (8.5) | 4.25 | < .001 |
| FM Group | 14.03 | (9.53) | 1.47 | .152 |
| Stimulation | 7.81 | (1.08) | 7.22 | < .001 |
|  | *σ^2^* | *(SD)* |  |  |
| *Random effects* | | | |  |
| Participant | 1052.9 | (32.45) | - | - |
| Participant x Stimulation | 29.7 | (5.45) | - | - |
| Linear mixed-effects model was fit by maximum likelihood, t-tests were calculated automatically using Satterthwaite approximations to degrees of freedom.  Coeff./σ^2^ – estimated coefficient/variance, SE/SD – estimated standard error/deviation. | | | | |

**Table 3. Model Summary for TSmax.t**

|  | *Coeff.* | *(SE)* | *t-value* | *p-value* |
| --- | --- | --- | --- | --- |
| *Fixed effects* | | | |  |
| Intercept | 2.64 | (1.28) | 2.05 | .048 |
| FM Group | 3.17 | (1.23) | 2.58 | .016 |
| Stimulation | .51 | (.09) | 5.46 | < .001 |
|  | *σ^2^* | *(SD)* |  |  |
| *Random effects* | | | |  |
| Participant | 16.96 | (4.12) | - | - |
| Participant x Stimulation | .09 | (.31) | - | - |
| Linear mixed-effects model was fit by maximum likelihood, t-tests were calculated automatically using Satterthwaite approximations to degrees of freedom.  Coeff./σ^2^ – estimated coefficient/variance, SE/SD – estimated standard error/deviation. | | | | |

Table 4. Model Summary for **TSm**

|  | *Coeff.* | *(SE)* | *t-value* | *p-value* |
| --- | --- | --- | --- | --- |
| *Fixed effects* | | | |  |
| Intercept | 5. | (1.19) | 4.24 | < .001 |
| FM Group | .41 | (.98) | .42 | .681 |
| Stimulation | .14 | (.08) | 1.8 | .089 |
|  | *σ^2^* | *(SD)* |  |  |
| *Random effects* | | | |  |
| Participant | 2.89 | (1.7) | - | - |
| Participant x Stimulation | 1.83 | (.01) | - | - |
| Linear mixed-effects model was fit by maximum likelihood, t-tests were calculated automatically using Satterthwaite approximations to degrees of freedom.  Coeff./σ^2^ – estimated coefficient/variance, SE/SD – estimated standard error/deviation. | | | | |

Table 5. Model Summary for **Aavg**

|  | *Coeff.* | *(SE)* | *t-value* | *p-value* |
| --- | --- | --- | --- | --- |
| *Fixed effects* | | | |  |
| Intercept | 45.18 | (9.36) | 4.83 | < .001 |
| FM Group | 20.04 | (10.6) | 1.89 | .069 |
| Stimulation | 8.15 | (.98) | 8.29 | < .001 |
|  | *σ^2^* | *(SD)* |  |  |
| *Random effects* | | | |  |
| Participant | 1314.5 | (36.26) | - | - |
| Participant x Stimulation | 24.3 | (4.93) | - | - |
| Linear mixed-effects model was fit by maximum likelihood, t-tests were calculated automatically using Satterthwaite approximations to degrees of freedom.  Coeff./σ^2^ – estimated coefficient/variance, SE/SD – estimated standard error/deviation. | | | | |

Table 6. Model Summary for **Amax.v**

|  | *Coeff.* | *(SE)* | *t-value* | *p-value* |
| --- | --- | --- | --- | --- |
| *Fixed effects* | | | |  |
| Intercept | 44.66 | (10.06) | 4.44 | < .001 |
| FM Group | 29.8 | (10.87) | 2.74 | .011 |
| Stimulation | 7.96 | (.97) | 8.23 | < .001 |
|  | *σ^2^* | *(SD)* |  |  |
| *Random effects* | | | |  |
| Participant | 1615.9 | (40.2) | - | - |
| Participant x Stimulation | 23.1 | (4.81) | - | - |
| Linear mixed-effects model was fit by maximum likelihood, t-tests were calculated automatically using Satterthwaite approximations to degrees of freedom.  Coeff./σ^2^ – estimated coefficient/variance, SE/SD – estimated standard error/deviation. | | | | |

Table 7. Model Summary for **Amax.t**

|  | *Coeff.* | *(SE)* | *t-value* | *p-value* |
| --- | --- | --- | --- | --- |
| *Fixed effects* | | | |  |
| Intercept | 12.66 | (4.44) | 2.85 | .007 |
| FM Group | 12.49 | (4.1) | 3.05 | .005 |
| Stimulation | 1.26 | (.37) | 3.41 | .006 |
|  | *σ^2^* | *(SD)* |  |  |
| *Random effects* | | | |  |
| Participant | 185.27 | (13.61) | - | - |
| Participant x Stimulation | 1.53 | (1.24) | - | - |
| Linear mixed-effects model was fit by maximum likelihood, t-tests were calculated automatically using Satterthwaite approximations to degrees of freedom.  Coeff./σ^2^ – estimated coefficient/variance, SE/SD – estimated standard error/deviation. | | | | |
